# Is my cough a cold or covid? A qualitative study of COVID-19 symptom recognition and attitudes towards testing in the UK

**DOI:** 10.1101/2021.05.28.21258022

**Authors:** Fiona Mowbray, Lisa Woodland, Louise E Smith, Richard Amlôt, G James Rubin

## Abstract

**Objective:** Key to reducing the spread of COVID-19 in the UK is increased use of the NHS Test and Trace (NHSTT) system. This study explored one of the main issues that determine whether people engage with NHSTT, how people understand symptoms that may indicate the presence of COVID-19 and that should trigger a request for a test.

**Methods:** In this qualitative study, a series of semi-structured telephone interviews were conducted with 40 people (21 members of the general population, 19 students). There was nearly an equal split between male and female participants in both populations. Data were collected between 30 November and 11 December 2020 and explored using thematic analysis. There was substantial similarity in responses for both populations so we combined our results and highlighted where differences were present.

**Results:** Participants generally had good knowledge of the main symptoms of COVID-19 (high temperature, new, persistent cough, anosmia) but had low confidence in their ability to differentiate them from symptoms of other illnesses. Attribution of symptoms to COVID-19 was most likely where the symptoms were severe, many symptoms were present, symptoms had lasted for some time and when perceived risk of exposure to infection was high due to previous contact with others. Participants felt encouraged to engage in testing where symptoms were present and had persisted for several days, though many had concerns about the safety of testing centres and the accuracy of test results. Students had mixed feelings about mass asymptomatic testing, seeing it as a way to access a more normal student experience, but also a potential waste of resources.

**Conclusions:** This study offers novel insights into how people attribute symptoms to COVID-19 and barriers and facilitators to engaging with testing. Participants had positive views of testing, but there is a need to improve not just recognition of each main symptom, but also understanding that even single, mild symptoms may necessitate a test rather than a “wait and see” approach, and to address concerns around test accuracy to increase testing uptake.

## Introduction

A central part of the UK Government’s COVID-19 response strategy, NHS Test and Trace (NHSTT) was launched in England in May 2020. This system aims to reduce the spread of SARS-CoV-2 by helping to identify and contain cases and their contacts. NHSTT relies on anyone experiencing one of the three symptoms described by the UK Government as the “main symptoms” of COVID-19 (high temperature; new, continuous cough; loss or change to sense of smell or taste) to self-isolate and request a test (1). People can choose to access a test either by travelling to a testing centre, or by having a test delivered to their home. If the test result is positive for SARS-CoV-2, self-isolation must continue for at least 10 days and the person is asked to provide details of any close contacts they have recently had with contacts inside and outside of the household. Contacts of cases are then contacted and must also self-isolate. Although a test, trace and isolate system is a useful tool to control the pandemic, success of the system is contingent upon high testing rates, high adherence to self-isolation and successful tracing of contacts (2, 3). A recent study found that only 51% of respondents in the UK were able to correctly identify COVID-19 symptoms, while only 18% of people who reported having symptoms of COVID-19 in the last 7 days requested a test. The most common reason for not requesting a test when symptomatic was thinking symptoms were not caused by COVID-19 (4). Uncertainty around how to interpret mild symptoms, and what symptom is experienced (e.g., whether it is a cough or fever) have also been suggested as reasons for either adopting a ‘wait and see’ attitude towards symptoms or not requesting a test at all (5, 6, 7). These potential delays to test seeking contribute some inefficiency to NHSTT and are particularly problematic as people are most infectious in the initial days after symptoms onset (8).

Another difficulty facing the testing system is that the proportion of asymptomatic COVID-19 infections has been found to be relatively high. A systematic review found that the proportion of asymptomatic infection among two general population studies at the time of testing was 20% and 75%, respectively (9). Conscious of the risk of asymptomatic viral transmission between students, staff and local communities that could come from the re-opening of university campuses (10), many UK universities rolled out regular mass asymptomatic testing during the Autumn 2020 term (11, 12). As with NHSTT, the success of mass asymptomatic testing relies on high levels of testing and adherence to self-isolation if a test result is positive. A recent study of wide-scale asymptomatic testing at one UK university found uptake to be low, dropping from 58% at the beginning of October 2020, to 5% by late-October. Key barriers to engaging in mass testing included concerns about the mental health impact of self-isolation and the impact on others if your test result is positive (13).

In this qualitative study we explored the key issues that underlie peoples’ engagement with NHSTT, specifically with regards to how people understand the symptoms that may indicate the presence of COVID-19 and that should trigger a request for a test. Given the focus in the UK on understanding how to prevent COVID-19 outbreaks in universities (e.g., 14) we assessed this in a sample of general members of the public and supplemented this with a separate sample of university students.

## Research Design and Methods

### Participant recruitment

We conducted 40 qualitative interviews with members of the general public (n = 21) and university students (n = 19) living in England. Recruitment was conducted by a specialist market research company and participants were selected by age, gender, region, ethnicity and place of residence. As we were particularly interested in exploring factors which may influence engagement with testing, we also recruited a subset of participants who had experienced cough, fever, or change / loss of sense of taste or smell within the last 7 days. Consent was obtained prior to data collection and ethical approval was granted by the Psychiatry, Nursing and Midwifery Research Ethics Subcommittee at King’s College London (LRS-20/21-21336:COVID-19).

### Interview procedure

For each participant, we conducted a semi-structured, one-to-one, telephone interview. These were conducted between 30 November 2020 and 11 December 2020 and lasted a mean of 38 minutes. It should be noted that the lockdown status within the UK changed during the study period. Initially there was a UK-wide lockdown (though schools were open), but this reverted to regional ‘tiered’ lockdown from 2 December 2020. Interviews were conducted by FM and LW, who are both female researchers with training and experience of qualitative methods in health research. Participants were not known to the researchers prior to the study, but they were informed about the purpose of the study and knew that the researchers were affiliated with King’s College London.

The interview guide was developed from relevant literature. Participants were asked a series of open questions to explore their perceptions and experiences of COVID-19-like symptoms and testing. This included questions about: their most recent cold or flu experience; knowledge and experience of COVID-19 symptoms; whether, when and why they would request a COVID-19 test; and any test experiences. A full interview guide is available in the supplementary materials. Each participant was compensated with a £40 e-gift card for taking part in the study.

### Data analysis

Interviews were audio recorded and transcribed verbatim by an external company. Each transcript underwent inductive thematic analysis (15), supported by use of the QSR NVivo 12 software, and was coded into emerging themes, which represented frequent patterns of meaning within the dataset. Coding followed the aims of the research, focusing on participants’ perceptions and experiences of COVID-19 symptoms and testing. Coding was done by FM, with the wider team frequently reviewing codes and advising on the development of themes. The final themes were agreed upon by the research team through discussion and consensus.

## Results

Participants from both the general population and student samples came from a range of regions across England and had a nearly equal split between female and male participants. The student population had a mean age of 20 (SD=1.7), while the mean age among the general population sample was 45 (SD=16.8). Students tended to live in shared accommodation (53%) or with parents (42%), while the general population tended to live alone (33%) or with a partner (48%). Both populations had a mix of participants who had or had not experienced symptoms in the previous 7 days. Full demographic details are presented in Table 1.

**Table 1.**
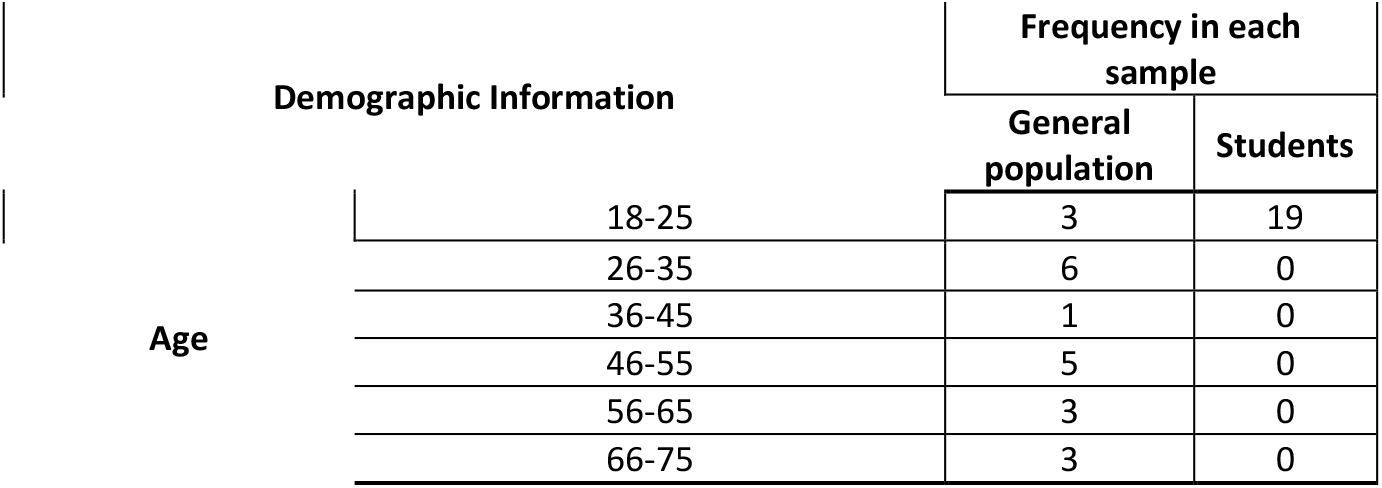
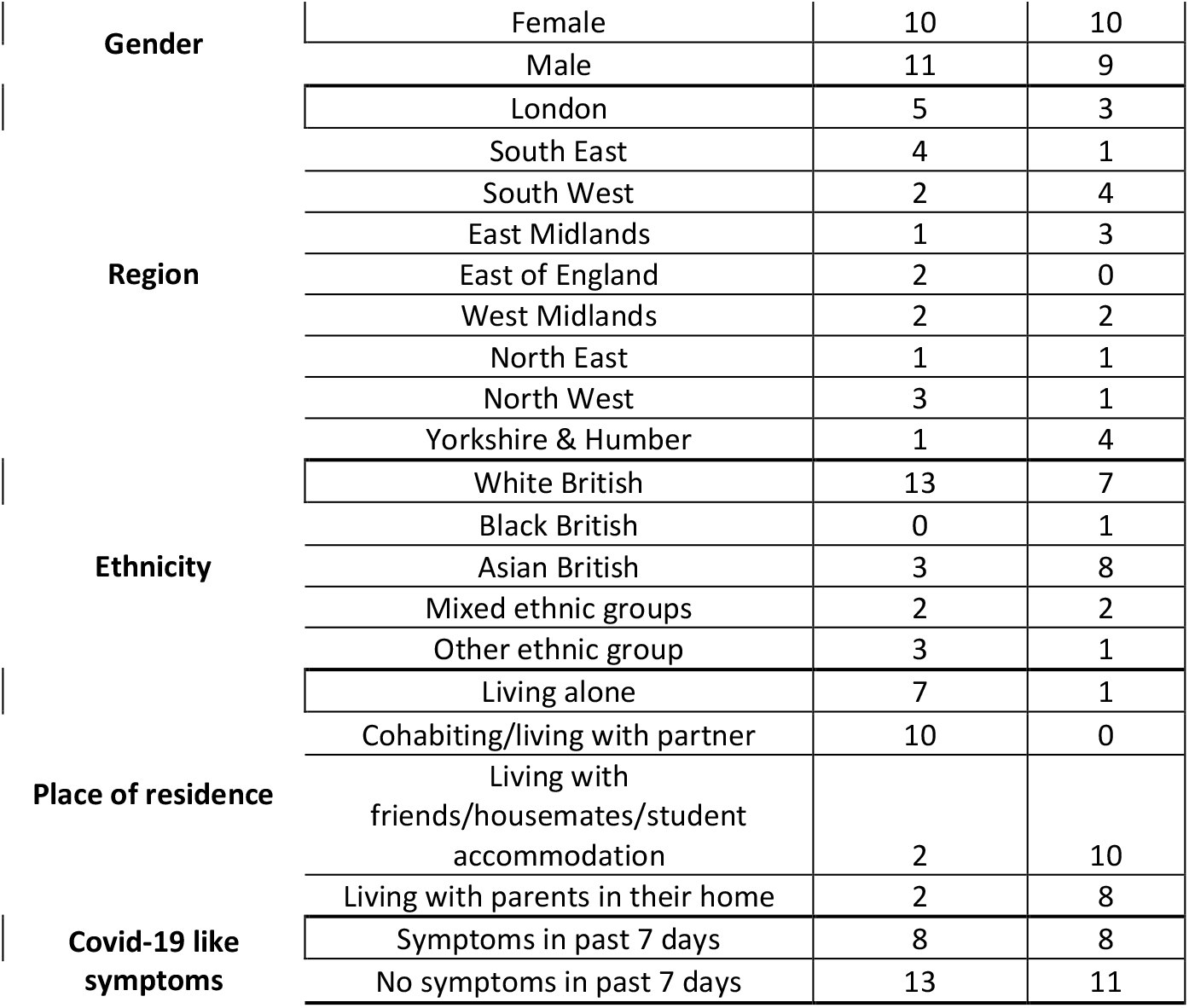
Participant demographic information

Following detailed thematic analysis, three interlinking main themes and ten subthemes were identified (Table 2). In line with the interview guide, participants all described their perceptions of COVID-19 symptoms, including how confident they felt about identifying symptoms and attributing these to COVID-19 versus another illness. This then led to discussions about when to seek a COVID-19 test and the specific facilitators and barriers to engaging with testing, as well as experiences of testing. Finally, participants spoke about the perceived impact of testing on their daily life, concerns about mass testing and frustrations about perceived low adherence of others to the guidance.

**Table 2.** Analytical framework for developing themes for participant perceptions and experiences of COVID-19 symptoms and testing

| **Theme 1: Factors affecting symptom attribution** |  |  |
| --- | --- | --- |
| **Subtheme** | **Definition** | **Example Quotes** |
| *Knowledge of COVID-19 symptoms* | Spontaneous and then prompted discussion about three main COVID-19 symptoms. Also includes details of experience of other people and potential 'other' symptoms of COVID-19. | *"If an individual has a cough, coupled with things like a fever...a cough generally has to be repetitive. A one-off cough isn't sufficient...also a loss of taste or smell." Participant 149, Student* |
| *Confidence identifying symptoms of COVID-19* | Discussion about confidence identifying symptoms of any illness and the challenges that COVID-19 symptom detection poses, particularly differentiating from symptoms of other illness. | *"I feel like there's a lot of crossover, I think the loss of taste and smell, that's quite distinct to Covid, but not everyone gets that. I think the cough and fever are more common, but you could experience that with a cold or flu."* |
|  |  | *Participant 111, General Population* |
| *Factors affecting attribution of symptoms to COVID-19* | Discussion about factors that would influence the attribution of symptoms to COVID-19. These were: perceived likelihood of contracting COVID-19 overall, symptom severity, amount and length, perceived exposure risk and engagement with mitigation strategies. | *"I think if I know that I haven't been in contact with anyone, I would think that it's probably just me being run down or tired. I wouldn't think it would be Covid if I wasn't in contact." Participant 101, General Population* |
| **Theme 2: Factors affecting test uptake** |  |  |
| **Subtheme** | **Definition** | **Example Quotes** |
| *Knowledge of test eligibility and access* | Participants' existing knowledge about who can or should request a test and how to access testing services. | *"As far as I'm aware, pretty much anyone is eligible if they have symptoms or if they have close contact with someone who has symptoms. I'm not sure about the latter part." Participant 105, General Population* |
| *Drivers to seeking a test* | Discussion about the reasons people have sought, or would seek, a test. These were: reassurance, a requirement for work or travel, the presence of symptoms. | *"I would only do it if the temperature was high and I had a continuous cough as well and I'd been out with my friends. If I had the symptoms then I would go and get tested, just to make sure that I was safe." Participant 146, Student* |
| *Barriers to engaging in testing* | Discussion about the factors that would discourage people from seeking testing. These were: concerns about safety of testing sites, discomfort of testing, concerns about test accuracy, stigma. | *"I was a little bit worried about what people might think if I'd caught it, like, what have you been doing to put yourself at risk? Have you not been following the guidelines?" Participant 109, General Population* |
| *Experience of testing* | Participant experiences of accessing and completing a COVID-19 test, as well as perceptions and experiences of NHSTT. | *"Every time I've been I think it's been a really efficient process, it's been really well structured, there's a lot of instructions from the people that work there. It does feel really safe when you're there." Participant 155, Student* |
| **Theme 3: Impact of testing on daily life and behaviour** |  |  |
| **Subtheme** | **Definition** | **Example Quotes** |
| *Experience of self-isolating* | Discussion about the impact on daily life and behaviour while waiting for a test result, including perceptions and experiences of self-isolating. | *"I hate just having to stay inside. It's a bit overwhelming at times, but at least I get on with everyone in my house. One of our friends would drop off our shopping and that was fine." Participant 148, Student* |
| *Perceptions about adherence of others to guidance* | Participant thoughts about whether others are following COVID-19 guidance and frustrations about those perceived to be flouting the rules. | *"It's being completely irresponsible on other people's health. Potentially someone can die if you don't self-isolate, so as hard as it is, it's my responsibility, it's everyone's responsibility to make* |
|  |  | *sure that everyone is safe.” Participant 103, General Population* |
| *Barriers and facilitators of mass asymptomatic testing* | Discussion from students about the testing offered by universities and the various benefits and concerns about this service. | *“I think it’s really beneficial because you wouldn’t want to give someone who’s slightly more at risk of COVID because you’d been out partying so I think a test is the best thing you can do.” Participant 158, Student* |

There was substantial similarity in responses for the general population and student samples. We have therefore combined the responses from both and highlighted where differences were present.

### Theme 1: Factors affecting symptom attribution

Participants across both groups generally demonstrated a good awareness of the three main, UK Government listed COVID-19 symptoms, often recalling at least 2 of these spontaneously and recognising others when prompted. Many participants also spoke about other symptoms that they or others had heard about or experienced, such as sore throat, gastrointestinal complaints, and fatigue, though participants were often unsure if these symptoms alone could indicate COVID-19. While most participants felt that they had a clear understanding of how each main symptom would present, there was some recognition that people may experience symptoms differently.

Overall, participants reported feeling confident in identifying symptoms of illness within themselves as they were either generally healthy and any difference would be noticeable, or they had another health condition which they felt made them more vigilant towards any changes. While some participants felt that they would be able to correctly attribute symptoms to COVID-19, the majority expressed concerns about their ability to determine whether a symptom was caused by COVID-19 versus another illness, though this varied by symptom. Anosmia (loss or change to taste or smell) was generally perceived to be more specific to COVID-19, making it easier to discern. A high temperature was not viewed as COVID-19 specific but was easy to detect and measure and there was an expectation that it would make you feel indisputably unwell. Many participants discussed the difficulty they would face when attempting to attribute a cough to COVID-19, due to its more variable nature:

> *“Coughing is the difficult one. You know if you’ve lost your sense of taste. We have a thermometer that I would use, but coughing…how frequently is frequently, and what does it actually mean to have a cough? Cough is such a standard symptom of so many things…it’s a more flexible kind of symptom*.*”* Participant 119, General Population

In general, there was an expectation among participants that experiencing a greater number and severity of symptoms increased the likelihood of them being caused by COVID-19 rather than another illness such as a cold or flu. Where symptoms were mild, attribution was much more difficult for participants and they displayed greater uncertainty in deciding what actions, if any, to take as a response to symptoms.

Symptom attribution was also dependent upon perceived risk, or likelihood of having been exposed to COVID-19. Generally, perceived risk appeared to be low among participants, but higher among those using public transport or going to work or university outside the home. Living in the same household as someone who used public transport or left home for work or other reasons also made participants feel at greater risk, even if they themselves were not engaging in these activities.

Participants reported that they felt more likely to attribute symptoms to COVID-19 if they had been in contact with others. Many participants also discussed beliefs that the greater the number of people they had been around and the closer or longer the contact, the more they felt likely that symptoms could be COVID-19:

> *“We all went out to this restaurant, but it wasn’t doing the social distancing…there were way too many people. My mum kept making jokes ‘If we’re going to catch it, we’re going to catch it here’. Three days later we started showing symptoms, so I was like, ‘We definitely have it from that restaurant*.*’ That’s what made us think, yes, it definitely is*.*”* Participant 157, Student

Conversely, most participants reported that if they had not been in close contact with others, they would be unlikely to attribute symptoms to COVID-19.

### Theme 2: Factors affecting test uptake

Most participants knew that anybody experiencing symptoms of COVID-19 was eligible to take a COVID-19 test and many listed certain groups who they perceived to be eligible regardless of symptoms, such as NHS or care home staff, teachers, and older adults. But despite this knowledge, participants generally displayed a lack of confidence about test eligibility, often questioning their understanding and discussing their perceptions that eligibility is ‘always changing’. Confidence in their ability to access a test when needed was high across all participants, with most mentioning searching online, the NHS website and some saying they would call the NHS 111 helpline or their GP if they were unsure. All students were aware that rapid lateral flow testing is available through their university and had received information about this process.

Participants all discussed engagement with symptomatic testing and for many this engagement would be driven by perceptions that it would be important to do if they had symptoms. They spoke about wanting to protect family and friends, or at-risk populations and feeling a ‘responsibility’ to ‘do the right thing’ to limit the spread of COVID-19. For some, an asymptomatic test was a requirement to travel, work, or visit someone and participants were willing to complete the test as a result. Several participants also mentioned being willing to take a test when symptomatic as they would be ‘curious’ to know if they had COVID-19 or perceived the test to provide ‘reassurance’ about their health. Although some participants said that they would seek a test at the first sign of any of the main symptoms, most said that they would ‘wait and see’ how symptoms developed over the course of several days, while also seeking out information on symptoms from official sources such as the NHS website. Generally, there was a feeling that people would wait for multiple symptoms to emerge before accessing a test, but anosmia would prompt some people to seek a test regardless of other symptoms as it was viewed as more ‘unusual’ or ‘novel’. A cough was also discussed by some as a driver to seek testing as it was sometimes perceived as posing a greater risk to others:

> *“…if you’re constantly coughing that’s most likely that you’re going to pass it on to other people or onto surface or objects. So yes, I think my first instinct would be, okay, I need to go and get a test*.*”* Participant 145, Student

Concerns about data privacy and missing university or work to attend a test rarely came up without prompting and were minimal for most participants. Often, any concerns were easily mitigated by seeking out information from trusted sources, for example, the NHS website. Similarly, although participants regularly mentioned the perceived discomfort of testing, this was viewed as something that just ‘needed to be done’ but would not discourage them from being tested. Some participants raised concerns about the safety of travelling to a testing site, particularly if they relied on public transport, but the perceived quicker result from a test site was preferable to waiting for a home test kit. Numerous participants spoke about the accuracy of test results, expressing concerns about possible false positives or negatives. Participants often reported low confidence in their perceived ability to perform the test correctly, worrying that this would lead to an inaccurate result:

> *“…it’s difficult to do it on your own because you’re always going to be hesitant to go deep enough. I think it’s probably best to get it done by people because there’s going to be a lot of inconclusive results and maybe it comes back as a false negative. I just feel like doing it on your own, there’s a lot of things that can go wrong*.*”* Participant 101, General Population

Many participants had concerns that testing may lead to stigma and feeling that others might perceive them as ‘risk taking’ by ‘not following the rules’. Students reported more varied perceptions of stigma. For some students, stigma was of minimal concern as it was deemed to be common for friends or housemates to engage with testing and a positive result was not unusual. Others had concerns about how friends and housemates might feel towards them if they tested positive and others needed to self-isolate as a result:

> *“Some of my flatmates have labs and face-to-face lectures and I’d feel bad that they’d have to miss that. Also, it’s nearly Christmas and a lot of them live away, they’d already booked tickets, so if I was to test positive they would miss that and I wasn’t looking forward to telling them, but luckily it was negative*.*”* Participant 156, Student

Experiences of test accessibility, organisation and speed of results were generally positive, however, those who had contact with the NHS Test and Trace system after testing positive provided negative feedback of this service, reporting that they had been called by contact tracers an ‘excessive’ number of times and given little useful support. One participant also reported having no contact from NHS Test and Trace despite testing positive. They found this ‘worrying’ and felt it eroded their trust in the system.

### Theme 3: Impact of testing on daily life and behaviour

Participants all discussed the process of waiting for test results, with most saying that if they were symptomatic, they would or did stay at home during this time to avoid spreading illness to others. Some said that if symptoms were mild or they ‘felt ok in themselves’ then they would still leave the house for essential shopping and to spend time outdoors in public spaces, although they would take measures such as mask wearing, physical distancing and hand washing to mitigate risk:

> *“I did try and self-isolate as often as I could, but I didn’t know definitely, so I still went to the supermarket, still went to different shops to get supplies*.*”* Participant 103, General Population

Similarly, those who had taken part in asymptomatic testing due to requirements for work or travel tended to report that they still went about their usual day-to-day activities while waiting for results, which they often received quickly if a rapid lateral flow test had been used. In the general population, self-isolation was perceived to be relatively easy as many people reported that it would not change much of their daily life while already in some level of lockdown and most felt they had someone who could provide support if needed. Those who had tested positive said that self-isolation had been somewhat difficult, but they generally reported experiencing high levels of support from family and friends. Students had mixed experiences of self-isolation with some saying it ‘was okay’ as their whole household had isolated together as a group, while others found it more difficult because they were alone in their room with little support available.

All participants expressed frustration towards people who did not engage in testing or self-isolation when deemed appropriate. They perceived this non-adherence to COVID-19 guidelines as ‘selfish’ and ‘dangerous’ behaviour that could put others at risk. Students often spoke about experiences where they felt that fellow students had behaved in ways that did not adhere to guidelines. They also had concerns about how students overall may be perceived negatively by the wider population as ‘not following the rules’, and ‘spreading COVID-19’:

> *“People probably think that students aren’t going to be affected by this so they don’t care, they’re just partying. Most students are not partying. So I think people just want a scapegoat*.*”* Participant 151, Student

Many students recognised that their age meant that COVID-19 was less likely to pose a serious risk to their health, and for some it felt like COVID-19 restrictions had an ‘unfair’ impact on the student demographic. However, there was often a feeling that their actions could negatively impact more vulnerable populations and students generally perceived a need to act ‘responsibly’.

Students were all aware of the testing available through their university, though perceptions about asymptomatic testing were mixed. For some students, testing was seen in a positive light, both as a way to ‘prove’ that they were behaving responsibly, but also because it could allow universities to keep operating in a more ‘normal’ fashion. Others were more sceptical about asymptomatic testing, suggesting that it was a ‘waste of resources’, may put people at risk by travelling to a site, and may force asymptomatic people to isolate and miss events that they otherwise would have been able to attend.

## Discussion

This study offers novel insights into how members of the public attribute symptoms to COVID-19, engage with testing and perceive the impact of both symptomatic and asymptomatic testing on daily life. Overall, most participants could name some symptoms of COVID-19 unprompted, but participants expressed low confidence in their ability to correctly attribute symptoms to COVID-19. Participants were more likely to both attribute symptoms to COVID-19 and to engage in testing where there was greater symptom severity, number of symptoms and symptom duration, or where perceived exposure was higher due to contact with others. Participants voiced concerns about the safety, accuracy, and potential impact of testing, but generally welcomed information from trusted sources to address these concerns. Individuals who had been through the NHSTT system generally reported positive experiences of accessing and completing a test, though there was an overall feeling that the NHSTT system needed improvement for people who tested positive.

Previous research indicates that knowledge of COVID-19 symptoms among the UK public is low, with only about half able to identify all three main symptoms (4, 16). In our study, while recognition of symptoms was reasonably good, it was clear that recognition alone was not always sufficient to trigger a decision to seek a test. Instead, the likelihood of seeking a test appeared to depend on the quality of the symptoms (number, severity, duration) and on the participant’s lay mental model of whether it was likely that they had been exposed to someone with COVID-19. Lay mental models are models of symptom appraisal, which highlight the interpretation of symptoms (rather than merely their detection) as being a crucial step in determining if and how people will respond to them (17). The finding has implications for communications about COVID-19 testing, which should emphasise not only what symptoms to watch out for, but also highlight that mild symptoms should trigger the same actions as severe symptoms, that a single symptom is just as important to check as multiple symptoms, and that ‘wait and see’ strategies or second-guessing the cause of a symptom are an example of risky behaviour. The finding that people base the decision to seek a test in part on whether they feel they have been at risk of infection corresponds with the results of studies in other contexts (18, 19) and may help to explain an apparent discrepancy in the UK data relating to testing uptake for COVID-19. For example, for the week of 5 to 11 November NHS TT identified 167,369 cases of COVID-19, largely via testing of symptomatic people (20). Over the same period, the UK’s Office for National Statistics estimated that there were 272,300 new cases of COVID-19 in England (95% credible interval: 240,100 to 308,700) (20). This suggests that roughly 60% of cases were detected by NHSTT. However, population surveys suggest that only around 20% of people with relevant symptoms during that period reported requesting a test (4). This discrepancy may suggest that the lay models used by people to judge whether their symptoms require a test or not are reasonably accurate.

Prior research has identified various logistical barriers to testing such as geographic, socioeconomic, and structural disparities in access (21, 22), while others report a lack of knowledge about how or where to access a test (16). In contrast, the current study found that participants reported high confidence in their ability to access testing if needed, both symptomatically through NHSTT for the general population and asymptomatically through universities for students. Those who had engaged with testing had found the system easily accessible and negative comments were largely reserved for the contact tracing element of the system. Instead, important barriers to testing included a lack of clarity about who should be tested and concerns about test accuracy. Confusion surrounding test eligibility appeared to come from information and guidelines that participants saw as often changing. Existing research has determined that a main reason for non-adherence to COVID-19 guidelines is “alert fatigue”, where the high volume and frequency of information received makes it difficult to follow guidelines (23). It is probable that this sense of overwhelm is also happening with information around test eligibility as people find the perceived everchanging criteria impedes their engagement with the system. Concerns about the accuracy of test results focussed on test sensitivity and self-administration of the test. Both those who have and have not engaged in testing felt worried about their ability to complete the swabbing part of testing effectively themselves, consistent with a recent study which found that 66% of participants preferred to be swabbed by a healthcare professional (24). There is a clear need to provide greater information and reassurance to people about test accuracy and to build greater self-efficacy. It may also be beneficial to highlight testing facilities where a ‘professional’ can conduct the swabbing, as trust in these staff is high and could encourage greater engagement with the system.

The importance of self-isolation both while waiting for test results and following a positive COVID-19 diagnosis was well understood by participants and willingness to self-isolate was high. Although many who had experience of self-isolation reported only partially adhering to self-isolation, they were generally engaging in sensible mitigation strategies based on perceptions about potential risk, which corresponds with findings from a recent study (25). Students also reported at least partial adherence to protective behaviours and as with previous studies (26), self-isolation was common, particularly when living in shared accommodation. Perceptions of mass asymptomatic testing were generally positive, but there was some hesitancy towards uptake. There were concerns that the tests may be inaccurate and that testing those without symptoms may be wasting finite resources that could be used for others who need them more. As with symptomatic testing, it would be beneficial to provide greater reassurance about the efficacy of rapid testing and clarify the importance of finding asymptomatic cases, particularly as they contribute to a quarter of all transmission (4). It would also be beneficial to clarify when lateral flow tests are most appropriate for use and that a positive lateral flow test should lead to a request for a PCR test. It is possible that asymptomatic testing may become more attractive as lockdown eases and people have the option to return to more social activities.

## Strengths and limitations

This qualitative study fills a knowledge gap of COVID-19 symptom attribution and testing engagement, using an in-depth analysis to generate understanding based upon the unique perspectives of the participants (27). It also helps to highlight key themes that should be considered in designing both future studies and communication materials, but has some limitations. Interviews were conducted in November and December 2020, ahead of the emergence of a more virulent strain of COVID-19 and the roll-out of the UK’s “roadmap” for emerging from pandemic restrictions, before the UK’s vaccination campaign, and across a period of changing restrictions. It is possible that views about symptom attribution and testing may have varied over time because of the overall changes to COVID-19 guidelines and perceived risk. Additionally, testing has become more available and accessible to all members of the public since data collection, for instance through rapid lateral flow tests, perhaps resulting in an alteration of the drivers and barriers to test engagement. This study relies on self-report data from participants about their perceptions, attitudes, and experiences and as a result, participants may have felt more motivated to report and advocate for behaviours they perceived to be more socially responsible. Furthermore, while some participants had personal experience of symptoms, engaging in testing, or a positive test result, many others had no, or only partial experience of these factors and were instead discussing their behavioural intentions. These intentions still provide important information to help us understand behaviour during the pandemic, but it is important to note that intentions are not always an accurate indicator of behaviour (28). Participants represented a wide range of socio-demographic characteristics, but it should be highlighted that all were proficient in English and had access to a telephone or internet connection.

## Conclusions

If policy makers wish to increase engagement with testing for COVID-19, there is a need to improve not just the recognition of symptoms among the public, but also their understanding of the need to seek testing for individual, mild symptoms without “waiting to see” if they resolve and without attempting to judge for themselves the likelihood of exposure having occurred.

## Data Availability

Data referred to in the manuscript can be accessed if requested by contacting the corresponding author.

## References

1. UK NHS. Get tested for coronavirus (COVID-19) (2021). https://www.nhs.uk/conditions/coronavirus-covid-19/testing/get-tested-for-coronavirus/ [Accessed May 18, 2021].

2. Kucharski AJ, Klepac P, Conlan AJK, Kissler SM, Tang ML, Fry H, Gog JR, Edmunds WJ. Effectiveness of isolation, testing, contact tracing, and physical distancing on reducing transmission of SARS-CoV-2 in different settings: A mathematical modelling study. Lancet Infect Dis (2020), 20:1151–1160. doi: 10.1016/S1473-3099(20)30457-6

3. Matukas LM, Dhalla IA, Laupacis A. Aggressively find, test, trace and isolate to beat COVID-19. Can Med Assoc J (2020), 192:40. doi: 10.1503/cmaj.202120

4. Smith LE, Potts HWW, Amlot R, Fear NT, Michie S, Rubin GJ. Adherence to the test, trace, and isolate system in the UK: results from 37 nationally representative surveys. BMJ (2021), 372. doi: 10.1136/bmj.n608

5. Fabella FE. Factors affecting willingness to be tested for COVID-19. SSRN [Preprint] (2020). Available at: https://papers.ssrn.com/sol3/papers.cfm?abstract_id=3670514 (Accessed May 4, 2021).

6. Hodson A, Woodland L, Smith LE, Rubin GJ. Parental perceptions of COVID-19-like illness in their children. Public Health (2021), 194:29–32. doi: 10.1016/j.puhe.2021.02.013.

7. Rubin GJ, Smith LE, Melendez-Torres GJ, Yardley L. Improving adherence to ‘test, trace and isolate’. J R Soc Med (2020), 113(9):335–338. doi: 10.1177/0141076820956824

8. Cevik M, Tate M, Lloyd O, Enrico Maraolo A, Schafers J, Ho A. SARS-CoV-2, SARS-CoV, and MERS-CoV viral load dynamics, duration of viral shedding, and infectiousness: a systematic review and meta-analysis. Lancet Microbe (2020), 2(1):E13–E22. doi: 10.1016/S2666-5247(20)30172-5

9. Yanes-Lane M, Winters N, Fregonese F, Bastos M, Perlman-Arrow S, Campbell JR, Menzies D. Proportion of asymptomatic infection among COVID-19 positive persons and their transmission potential: A systematic review and meta-analysis. PLoS ONE (2020), 15(11):e0241536. doi: 10.1371/journal.pone.0241536

10. Yamey G, Walensky RP. COVID-19: Re-opening universities is high risk. BMJ (2020), 370:m3365. doi: 10.1136/bmj.m3365

11. Berger Gillam T, Cole J, Gharbi K, Angiolini E, Barker T, Bickerton P, Brabbs T, Chin J, Coen E, Cossey S, Davey R, Davidson R, Durrant A, Edwards D, Hall N, Henderson S, Hitchcock M, Irish N, Lipscombe J, Jones G, Parr G, Rushworth S, Shearer N, Smith R, Steel N. Norwich COVID-19 testing initiative pilot: Evaluating the feasibility of asymptomatic testing on a university campus. J Public Health (2020), 43(1):82–88. doi: 10.1093/pubmed/fdaa194

12. Blake H, Corner J, Cirelli C, Hassard J, Briggs L, Daly JM, Bennett M, Chappell JG, Fairclough L, McClure CP, Tarr A, Tighe P, Favier A, Irving W, Ball J. Perceptions and experiences of the University of Nottingham pilot SARS-CoV-2 asymptomatic testing service: A mixed methods study. Int J Environ Res Public Health (2020), 18(1):188. doi: 10.3390/ijerph18010188

13. Blake H, Knight H, Jia R, Corner J, Morling JR, Denning C, Ball JK, Bolton K, Figueredo G, Morris DE, Tighe P, Mendez Villalon A, Ayling K, Vedhara K. Students’ views towards SARS-CoV-2 mass asymptomatic testing, social distancing and self-isolation in a university setting during the COVID-19 pandemic: A qualitative study. Int J Environ Res Public Health (2021), 18:4182. doi: 10.3390/ijerph18084182

14. HM Government. Principles for managing SARS-CoV-2 transmission associated with higher education (2020). https://assets.publishing.service.gov.uk/government/uploads/system/uploads/attachment_data/file/914978/S0728_Principles_for_Managing_SARS-CoV-2_Transmission_Associated_with_Higher_Education_.pdf [Accessed May 16, 2021].

15. Joffe H & Yardley L. “Content and thematic analysis,”. In: Marks DF & Yardley L, editors. Research methods for clinical and health psychology. London: Sage (2004).

16. Graham MS, May A, Varsavsky T, Sudre CH, Murray B, Klaser K, Antonelli M, Canas LS, Molteni E, Modat M, Jorge Cardoso M, Drew DA, Nguyen LH, Rader B, Hu C, Capdevila J, Hammers A, Chan AT, Wolf J, Brownstein JS, Spector TD, Ourselin S, Steves CJ, Astley CM. Knowledge barriers in the symptomatic-COVID-19 testing programme in the UK: an observational study. MedRxiv [Preprint] (2021). Available at: https://www.medrxiv.org/content/10.1101/2021.03.16.21253719v2.full (Accessed May 10, 2021).

17. Whitaker, SE, Scott SE, Wardle J. Applying symptom appraisal models to understand sociodemographic differences in responses to possible cancer symptoms: a research agenda. Br J Cancer (2015), 112:S27–S34.

18. Rubin GJ, Page L, Morgan O, Pinder RJ, Riley P, Hatch S, Maguire H, Catchpole M, Simpson J & Wessely S. Public information needs after the poisoning of Alexander Litvinenko with polonium-210 in London: Cross sectional telephone survey and qualitative analysis. BMJ (2007), 335(7630):1143–6. doi:10.1136/bmj.39367.455243.BE

19. Rubin GJ, Amlôt R, Carter H, Large S, Wessely S, Page L. Reassuring and managing patients with concerns about swine flu: Qualitative interviews with callers to NHS Direct. BMC Public Health (2010), 10. doi:10.1186/1471-2458-10-451

20. Department of Health & Social Care. Weekly statistics for NHSTest and Trace (England) and coronavirus testing (UK): 5 November to 11 November (2020). https://www.gov.uk/government/publications/nhs-test-and-trace-england-and-coronavirus-testing-uk-statistics-5-november-to-11-november/weekly-statistics-for-nhs-test-and-trace-england-and-coronavirus-testing-uk-5-november-to-11-november#fnref:1. [Accessed 30 November 2020].

21. Rader B, Astley CM, Sy KTL, Sewalk K, Hswen Y, Brownstein JS, Kraemer MUG. Geographic access to United States SARS-CoV-2 testing sites highlights healthcare disparities and may bias transmission estimates. J Travel Med (2020), 27. doi: 10.1093/jtm/taaa076

22. Thunstrom L, Ashworth M, Shogren JF, Newbold S, Finnoff D. Testing for COVID-19: willful ignorance or selfless behaviour? Behav Public Policy (2021), 5(2):135–152. doi: 10.1017/bpp.2020.15

23. Williams SN, Armitage CJ, Tampe T, Dienes KD. Public perceptions of non-adherence to COVID-19 measures by self and others in the United Kingdom. MedRxiv [Preprint] (2020). Available at: https://www.medrxiv.org/content/10.1101/2020.11.17.20233486v1.full#:~:text=In%20the%20UK%2C%20longitudinal%20public,during%20the%20country’s%20’second%20wave’ (Accessed May 14, 2021).

24. Wurstle S, Spinner CD, Voit F, Hoffman D, Hering S, Weidlich S, Schneider J, Zink A, Treiber M, Iakoubov R, Schmid RM, Protzer U, Erber J. Self-sampling versus health care professional-guided swab collection for SARS-CoV-2 testing. Infection (2021). doi: 10.1007/s15010-021-01614-9

25. Denford S, Morton KS, Lambert H, Zhang J, Smith LE, Rubin GJ, Cai S, Zhang T, Robin C, Lasseter G, Hickman M, Oliver I, Yardley L. Understanding patterns of adherence to COVID-19 mitigation measures: a qualitative interview study. J Public Health (2021). doi: 10.1093/pubmed/fdab005

26. Nixon E, Trickey A, Christensen H, Finn A, Thomas A, Relton C, Montgomery C, Hemani G, Metz J, Walker JG, Turner K, Kwiatkowska R, Sauchelli S, Danon L, Brooks-Pollock E. Contacts and behaviours of university students during the COVID-19 pandemic at the start of the 2020/21 academic year. MedRxiv [Preprint] (2020). Available at: https://www.medrxiv.org/content/10.1101/2020.12.09.20246421v1.full (Accessed May 14, 2021).

27. Teti M, Schatz E, Liebenberg L. Methods in the time of COVID-19: the vital role of qualitative inquiries. Int J Qual Methods (2020), 19:1–5. doi: 10.1177/1609406920920962

28. Webb TL, Sheeran P. Does changing behavioral intentions engender behaviour change? A meta-analysis of the experimental evidence. Psyc Bull (2006), 132(2):249–28. doi:10.1037/0033-2909.132.2.249

