## Supplementary Material 1 for "Is my cough a cold or covid? A qualitative study of COVID-19 symptom recognition and attitudes towards testing in the UK"

**Interview Topic Guide**

### Knowledge, perceptions, experience of symptoms

Can you tell me a bit about the last time you had a cold or the flu?

- When was that?
- Were your symptoms mild or severe or somewhere in between?
- How did you manage your symptoms?

Can you tell me about the symptoms of Covid-19? *(see what if any symptoms they cover, only prompt if they don't mention them)*

- Fever: tell me about what you consider to be a fever? How do you measure it?
- Cough: tell me about what you think counts as a cough? How long, how frequent, etc.
- Loss or change to sense of smell/taste: tell me what this means to you? What kind of change?
- Any other symptoms (e.g. sore throat, sneezing, runny nose, headache): What do these mean to you? What would you do about them?

How confident do you feel in identifying symptoms in yourself/your child? If not confident, what might make it easier?

If you have more than 1 child, do you think you would react to symptoms differently depending on which child displayed symptoms?

How confident do you feel being able to tell the difference between Covid-19 and a cold or other illness in yourself/your child? Can you tell me about how you're making that decision?

When would you still go out/send your child to school? What made that important to you?

In what way would how you act on symptoms that you think might be Covid-19 differ from how you would act on symptoms that might not be caused by Covid-19 / have acted on symptoms in the past?

How likely do you feel you/your child are to get Covid-19? Can you tell me about why you do or do not feel at risk?

Have you/your child experienced any of these symptoms (fever, cough, loss/change in taste/smell) in the last 7 days?

- If so, can you tell me step by step about the symptom/s and the experience? Combination of symptoms or just 1? How severe were symptoms?
- What if anything did you do about the symptom(s)? Stay off school/work?
- Did you search for information about the symptom(s)? If no, why not? If yes, what information and where did you look? Was it helpful?

### Knowledge, perceptions, experience of testing

Who do you think is eligible to get tested for Covid-19? What makes someone eligible?

When would you ask for a Covid-19 test for yourself/your child?

- How long after symptoms start?
- How many symptoms? Just 1? Combination?

- How severe are the symptoms?

Have you/your child ever had a Covid-19 test?

- If yes:
  - Can you tell me a bit about this experience? What led you to getting a test?
  - Did you have any concerns about getting tested? (data privacy, social stigma or reaction of friends/family/colleagues, missing work/school, loss of income, safety concerns, effectiveness of the test, etc.)?
  - Is there anything that would ever stop you getting a test? Why or why not?
  - Did you/they have any symptoms? If so, what were your/their symptom(s)?
  - When did you seek the test? How long after symptom(s)?
  - How did you get the test? At home, test centre, etc. Could anything make the process easier? If you went to a test centre, did you feel safe?
  - What was the test like? Can you tell me about what you had to do?
  - What happened after you finished the test?
  - How did you get your results?
  - How long were you waiting for your results? Did you do anything differently while you were waiting for your results to come through? How did you feel during this time?
  - Did you feel that people treated you any differently after learning that you'd had a test for coronavirus (regardless of the outcome)?
  - If positive, can you tell me about whether you self-isolated and what this was like? What made it more difficult? What might make it easier? If did not isolate, why not?
  - If positive, did you get in touch with other people to let them know you had tested positive? Did / do they treat you any differently?
  - Were you contacted by Test and Trace? If so, can you tell me a bit about this experience?
- If no:
  - Can you tell me about how you would go about getting a Covid-19 test?
  - Where would you go for information? What information would you want/need?
  - Would you want to be tested? If no, why not? What concerns do you have (data privacy, social stigma or reaction of friends/family/colleagues, missing work/school, loss of income, safety concerns, effectiveness of the test, etc.)?
  - How would you feel about doing the test (would it hurt/be uncomfortable, confident able to do it yourself/for your child)?
  - Would you do anything differently after being tested for coronavirus?
  - Do you think people would treat you differently if you were tested for coronavirus?
    - If positive / If negative
  - Can you tell me about what you would do if the test came back positive?
  - What impact would a positive result have on you/your child's life?

Has anyone you know had a Covid-19 test? What if anything did they tell you about the experience?

What do you think would happen if someone had a positive test result, but did not self-isolate?

What if anything would encourage you to take a test?

What information would you want about testing? Where/who do you want the information coming from? Why? Anyone you do not want information from? Why?

Students only:

Has your university provided any information about testing? If so, can you tell me a bit about this?
